# Circulating fibrocytes can identify people with a poor prognosis in idiopathic pulmonary fibrosis

**DOI:** 10.1101/2020.06.05.20123406

**Authors:** Iain D. Stewart, Henry Nanji, Grazziela Figueredo, William A. Fahy, Toby M. Maher, Antje J. Ask, Shyam Maharaj, Kjetil Ask, Martin Kolb, Gisli R. Jenkins

## Abstract

**Objective:** Circulating fibrocytes are elevated in idiopathic pulmonary fibrosis, but the relationship between fibrocyte level with lung function decline and outcomes is lacking replication in prospective clinical study. We aim to validate the utility of circulating fibrocyte levels as a prognostic biomarker in idiopathic pulmonary fibrosis.

**Methods:** We tested associations between circulating fibrocyte levels, mortality, disease progression and longitudinal lung function in a well-defined prospective observational study of pulmonary fibrosis (PROFILE; NCT01134822). A subset of recruited participants had blood samples processed for fibrocyte measurement, with flow cytometry based on CD45 and collagen-I gating. Associations were tested using univariable and multivariable generalised linear models. Mortality data were subsequently combined with an independent cohort in a mixed-effect multilevel analysis.

**Results:** In 102 participants with idiopathic pulmonary fibrosis, an empirically defined cutpoint of 2.22% was associated with a greater risk of overall mortality in adjusted analysis (Hazard Ratio 2.24 95% CI 1.06-4.72). A 2.5 fold greater risk of mortality was supported in a pooled analysis with a historic cohort for a larger sample of 162 participants of idiopathic pulmonary fibrosis, specifically (Hazard Ratio 2.49 95% CI 2.41-2.56). A previously defined mortality risk threshold of 5% circulating fibrocytes was not reproducible in this cohort, circulating fibrocytes were not significantly elevated above non-specific interstitial pneumonia or healthy controls.. We found no association of fibrocytes with lung function or disease progression.

**Conclusions:** In a prospective clinical cohort of idiopathic pulmonary fibrosis, circulating fibrocytes of 2.22% or above are associated with greater mortality, but do not associate with disease related decline in lung function.

**What is the key question?:** Can circulating fibrocytes provide reproducible prognostic biomarker value in fibrotic lung disease?

**What is the bottom line?:** Greater proportions of fibrocytes isolated from circulating leukocyte populations are associated with an increase risk of mortality in idiopathic pulmonary fibrosis, but no association was observed with disease related decline in lung function.

**Why read on?:** We present associations and limitations of circulating fibrocytes in the largest sample of individuals with pulmonary fibrosis, recruited into a prospective observational study, and replicate prognositic insights from a historic cohort.

## Introduction

Pulmonary fibrosis is the process of progressive scarring of lung tissue due to dysregulation of extracellular matrix turnover at the site of wound healing, the hallmark feature of a number of respiratory diseases.^1^ Idiopathic pulmonary fibrosis (IPF) is a particularly severe and well-characterised type of pulmonary fibrosis with a 3-year median survival and an increasing incidence globally.^2^ Individuals with IPF have varying degrees of fibrosis at presentation, and heterogeneneous progression of their illness. Understanding the mechanisms underpinning this variability may provide new therapeutic avenues or biomarkers of disease.

Fibrocytes are circulating mesenchymal progenitor cells that differentiate into tissue specific fibroblasts and contribute to multiple wound healing processes, including secretion of inflammatory cytokines, contractile wound closure, and promotion of angiogenesis.^3^ The mesenchymal properties of the fibrocyte enables tissue invasion where it is thought they have pathogenic roles in fibrosis through transdifferentiation and accumulation of activated fibroblasts without resolution, or via paracrine signalling to resident cell types ultimately leading to production of dysregulated collagens through TGF-β mediated pathways.^4^

The role of fibrocytes in IPF remains controversial. Studies in transgenic mouse models have demonstrated that fibrocytes are not a necessary source of collagen-I and do not contribute to aberrant deposition in lung fibrosis.^5^ However, a clinical study observed higher fibrocyte levels during IPF exacerbation and a significant association with overall mortality at a threshold of 5%^6^ Thus, the contribution of fibrocytes to the pathogenesis of progressive pulmonary fibrosis remains unclear and clinical observations require validation in independent cohorts.

We utilised the PROFILE study, a prospective multi-centre longitudinal cohort of patients with progressive fibrotic Interstitial Lung Diseases including IPF and fibrotic NSIP,^7 8^ to validate whether circulating fibrocytes are reliably associated with key clinical outcomes in idiopathic pulmonary fibrosis, including overall mortality, disease progression and lung function decline.

## Methods

### Participants and Study Design

Individuals with IPF or idiopathic NSIP confirmed by multidisciplinary team assessment were recruited to the PROFILE study as previously described.^7 8^ Blood samples were collected at baseline and six months and processed for circulating fibrocyte numbers in 119 PROFILE participants; samples from age-matched healthy controls were also collected for comparison. Individuals in the PROFILE study had demographic data and lung function recorded with twelve months of clinical follow up available. Disease progression was defined as a 10% relative decline in FVC, or death, within 12 months. Survival data were censored at 31 March 2016. Data were subsequently pooled with independent circulating fibrocyte data from individual IPF patients (Moeller *et al*. 2009),^6^ diagnosed using the same standards.^9^

### Fibrocyte Processing and Measurement

Fresh blood samples were centrifuged in Accuspin System Histopaque-1077 tubes at lOOOx g for 10 mins. The mononuclear cell layer was transferred and resuspended before storage in liquid nitrogen, and shipped as a single batch on dry-ice. Fibrocytes were measured as described in Moeller et al.^6^ Briefly, cells were first immunostained for the surface antigen CD45 (CD45-PerCP, #557513; BD Biosciences) or isotype control antibody (IgG_1_, #555751; BD Biosciences), and permeabilized for detection of intracellular antigens. Cells were incubated with specific rabbit anti-human collagen-1 antibody (#6004011030.1 Rockland Immunochemicals) or IgG isotype control antibody (#AB105C; R&D Systems) followed with secondary antibody conjugated to Alexa Fluor 488 (#A20981; Invitrogen). Flow cytometry was performed with FACSDiva software (BD Biosciences). All data were analyzed with FlowJo software (Tree Star, Inc., Ashland, OR). The negative threshold for CD45 was set at 0.5%, using unstained cells, and all subsequent samples were gated for the CD45+ region. Cells gated for CD45 were analyzed for collagen-1 expression, with negative control thresholds set at 0.5%, using a matched IgG isotype control. Specific staining for collagen-1 was determined as an increase in positive events over this threshold. Fibrocyte levels are defined as the percentage of circulating fibrocytes in total circulating leukocytes. Changes in paired fibrocyte levels at baseline and six months were compared where available (Supplementary Table 1). There were no measurable or statistically significant differences between paired values, we use the first chronologically occurring fibrocyte measurement for all individuals to avoid missingness.

### Statistical Analysis

Fibrocyte levels were compared between IPF, NSIP and healthy controls using nonparametric tests, addressing disease status. Kaplan-Meier estimates of potential follow-up (KM-PF) are used to describe distribution of follow-up time. Kaplan-Meier survival analyses were performed at a threshold of 5%,^6^ assessed by non-parametric log-rank test. An empirical cutpoint was defined in IPF using the Youden method. Cox proportional hazard models assessed univariable and adjusted overall mortality risk for a 5% threshold in fibrocyte levels, and based on the empirical cutpoint, restricted to IPF diagnoses. Hazard ratios are presented with 95% confidence intervals adjusted for robust standard errors, and with adjustment for *a priori* confounders including percent predicted baseline FVC and DLco, age, gender, ever smoker status and steroid immunosuppression background therapy. Cox proportional hazard models in the mixed effect pooled analysis were fit with a Weibull distribution and using a random intercept at the study level.

Associations with lung function metrics FVC, DLCO and FEV1/FVC ratio at baseline and change over time were assessed according to fibrocyte level in generalised linear models. Differences in fibrocyte level according to disease progression status was assessed by t-test. Statistical significance was defined as p<0.05 throughout. All analyses were performed with Stata 16.0 (StataCorp TM).

## Results

In total, 102 (85.7%) PROFILE participants sampled for fibrocyte measurement were diagnosed with IPF and 17 (14.3%) participants were diagnosed with NSIP. Baseline characteristics of NSIP and IPF patients were similar, although there was a greater proportion of NSIP participants on steroid immunosuppression (Table 1). IPF diagnosed participants had a median 73 years of age (IQR 68-79), 75% male and 78% ever smokers. FVC was 82.8% (±20.7) of predicted value on average, whilst DLCO was 48.5% (±16.7) of predicted value. Lung function was slightly higher in sampled participants compared to those not sampled for fibrocyte measurement, and were otherwise representative (Supplementary Table 2).

**Table 1.**
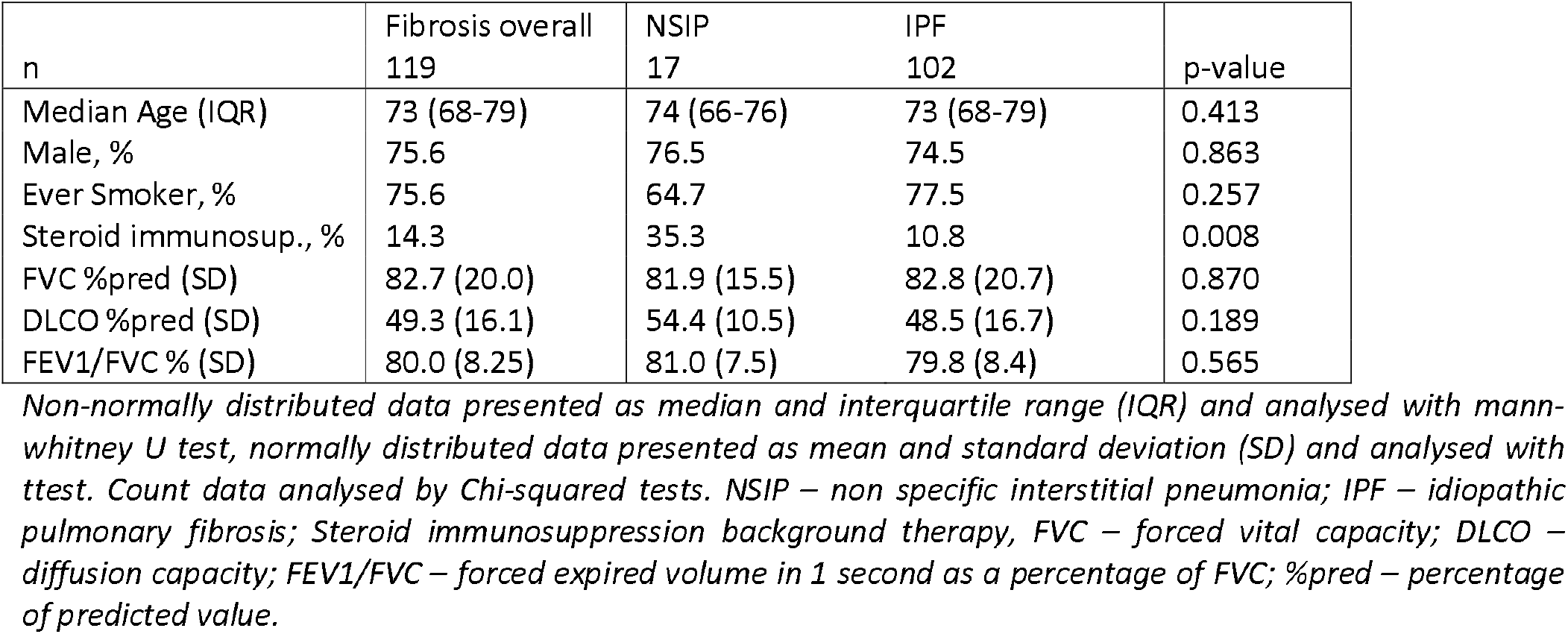
Baseline characteristics according to disease status.

**Table 2.** Association of lung function indices and fibrocyte level.

|  | n | Coef. | 95%CI | p-value | Coef.* | 95% CI | p-value |
| --- | --- | --- | --- | --- | --- | --- | --- |
| FVC %pred | 119 | -0.029 | (-0.088; 0.029) | 0.325 | -0.024 | (-0.084; 0.037) | 0.443 |
| DLCO %pred | 105 | -0.025 | (-0.101; 0.049) | 0.498 | -0.031 | (-0.107; 0.045) | 0.423 |
| FEV1/FVC | 119 | 0.057 | (-0.085; 0.120) | 0.430 | 0.030 | (-0.118; 0.179) | 0.690 |

*\*adjusted for gender, age, ILD subtype, and smoking*

The percentage of circulating fibrocytes was highly variable across all participant groups. Median circulating fibrocyte level was higher in NSIP (3.84%) than healthy controls (1.61%), whilst levels were highest in IPF (4.49%), but these were not significantly different (Figure 1). The range of fibrocyte levels extended furthest in IPF (38.6%), compared with NSIP (24.1%) and controls (16.6%).

**Figure 1.**
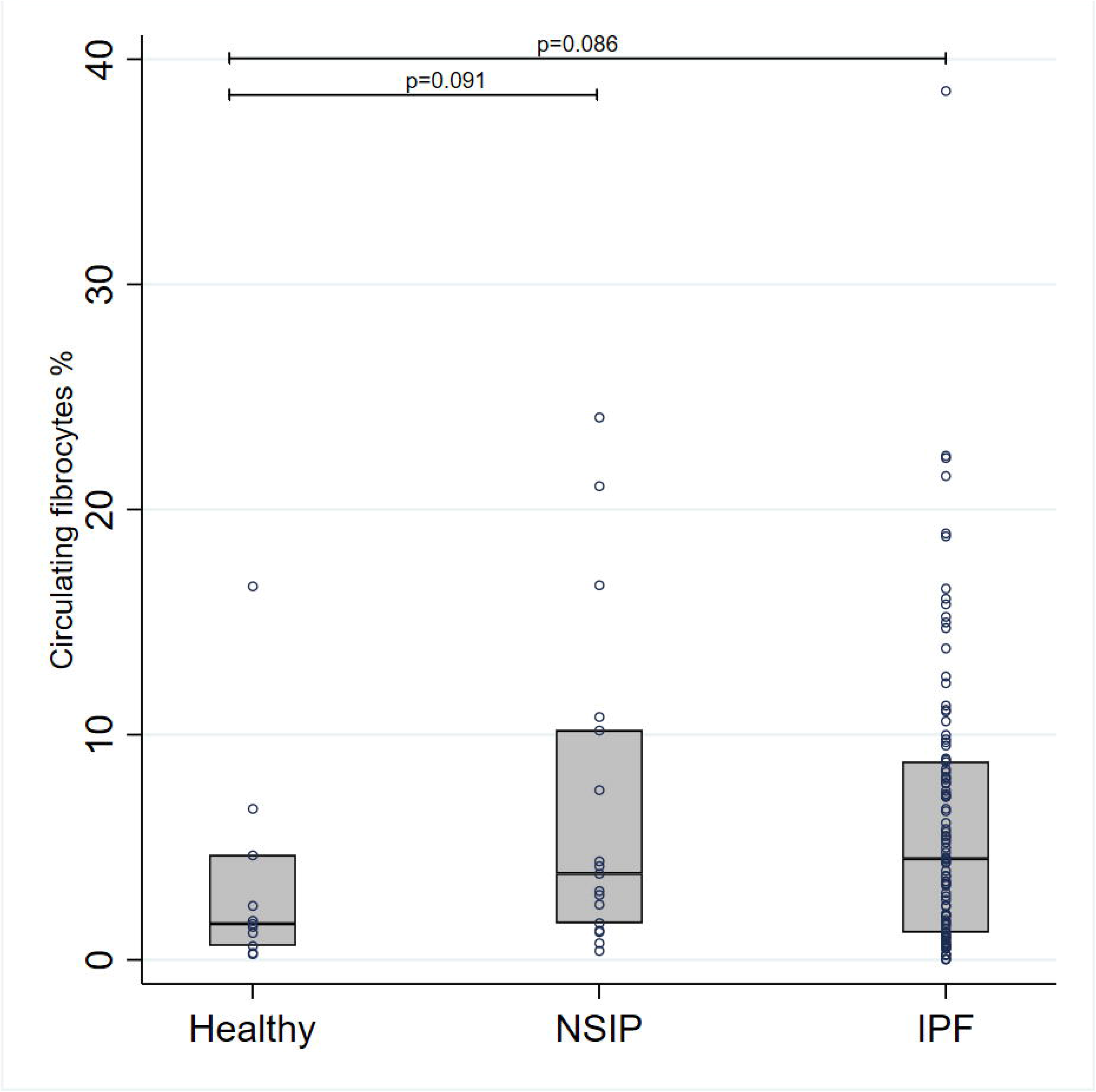
Median circulating fibrocyte level by disease status. Fibrocyte levels (%) plotted according to disease diagnosis of NSIP or IPF, or from healthy age matched control. Median and interquartile range are presented for healthy (1.61 IQR: 0.63-4.66), NSIP (3.84 IQR: 1.64-10.20), and IPF (4.49 IQR 1.22-8.80). P-values provided for the difference between Healthy and NSIP (p=0.091), the difference between Healthy and IPF (p=0.086).

**Figure 2.**
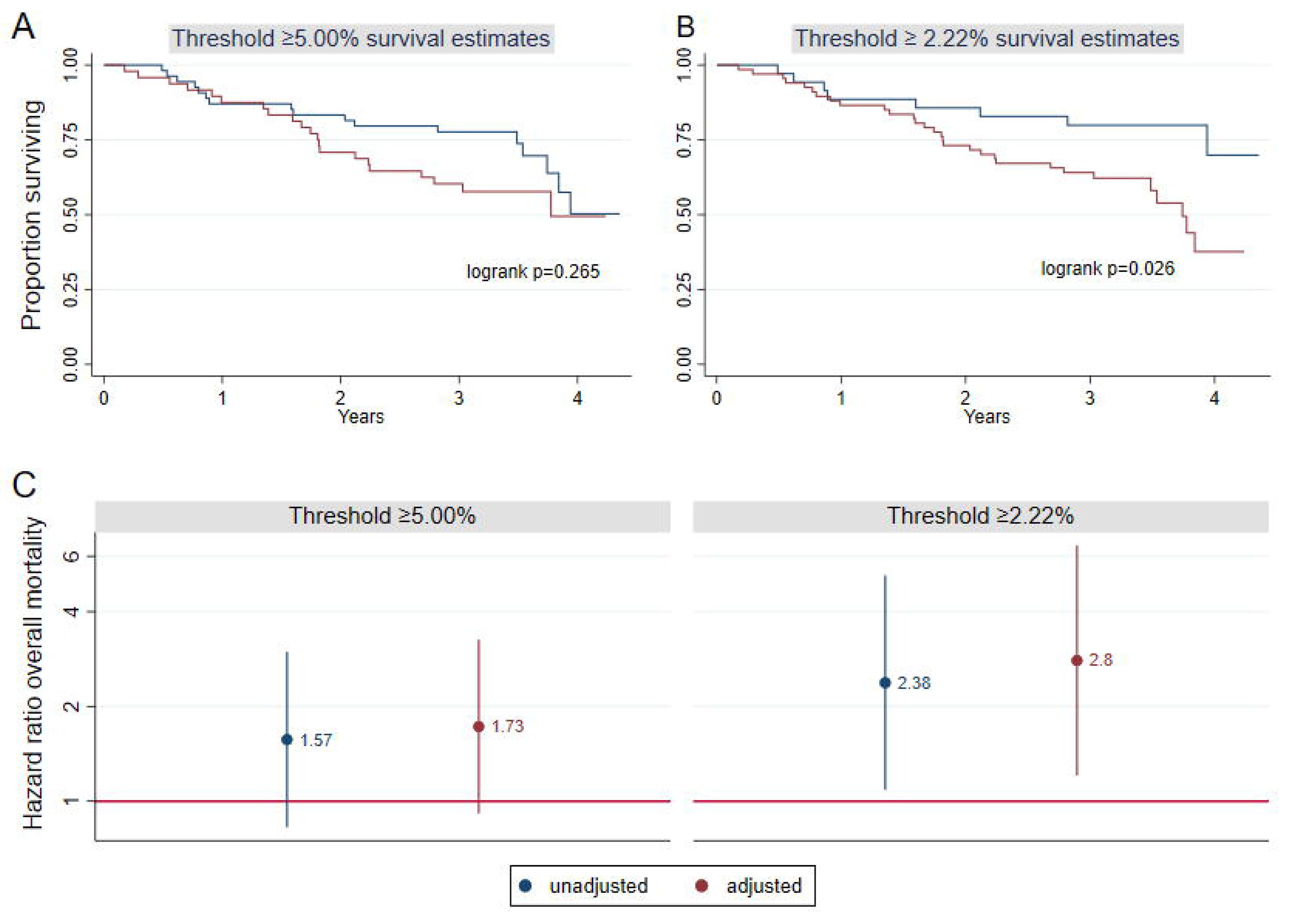
Survival curve and mortality risk for threshold of circulating fibrocyte level in IPF. Proportion surviving plotted against follow up time in years for threshold defined at (A) 5.00% circulating fibrocytes and (B) 2.22%, threshold survival compared with logrank test; below threshold blue line, above threshold red line. (C) Unadjusted hazard ratio estimates and estimates adjusted for age, gender, baseline percent predicted FVC and steroid immunosuppression, presented with 95% confidence intervals.

In survival analyses, participants were followed up for a median 38.1 months (IQR 25.8–44.1). Univariable analysis of fibrocyte level in participants with IPF indicated that a 5.0% threshold was not associated with mortality in univariable or adjusted analysis (HR 1.57 95%CI 0.83-2.98; HR 1.73 95%CI 0.91-3.26, respectively). Based on maximal sensitivity and specicity (Supplementary Figure 1), an empirically derived threshold of 2.22% was significantly associated with a greater than 2-fold risk of overall mortality (HR 2.38 95%CI 1.09-5.21), which remain significant following adjustment (HR 2.80 95%CI 1.21-6.49, Figure 3).

**Figure 3.**
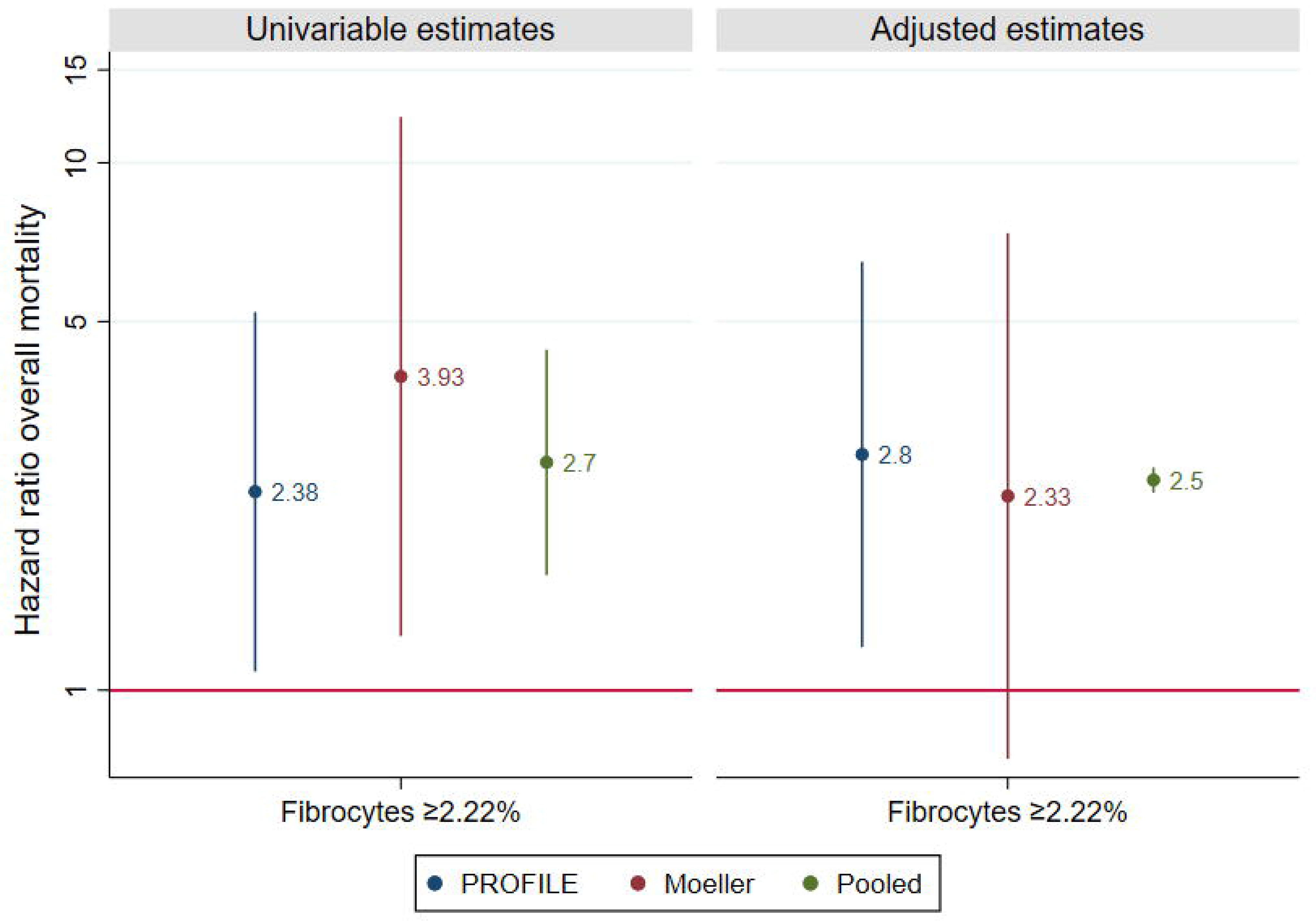
Hazard ratio of fibrocyte threshold in IPF pooled analysis. Association of dichotomised fibrocyte level at 2.22% threshold with overall risk of mortality. Hazard ratios presented with 95%CI in unadjusted analysis, and analysis adjusted for age, gender, baseline percent predicted FVC and steroid immunosuppression. Analysis performed in PROFILE participants, Moeller et al participants, and pooled in mixed effect multilevel model.

To maximise power in evaluating the prognostic capacity of the empirical cutpoint of circulating fibrocytes, and to compare estimates with a historical, independent cohort, we performed pooled analysis of PROFILE data with data from Moeller *et al*. (2009) for people with IPF specifically (usual interstitial pneumonia pattern). Unadjusted estimates were significant in PROFILE, Moeller *et al*. and overall (Figure 4, Supplementary Table 3). Significance was not observed in adjusted estimates for data from Moeller *et al*. In a multilevel analysis adjusted for gender, age, baseline FVC and background immunosuppression therapy, participants with a circulating fibrocyte level ≥2.22% were at a 2.5-fold greater risk of mortality (HR 2.5 95%CI 2.37-2.64).

**Figure 4.**
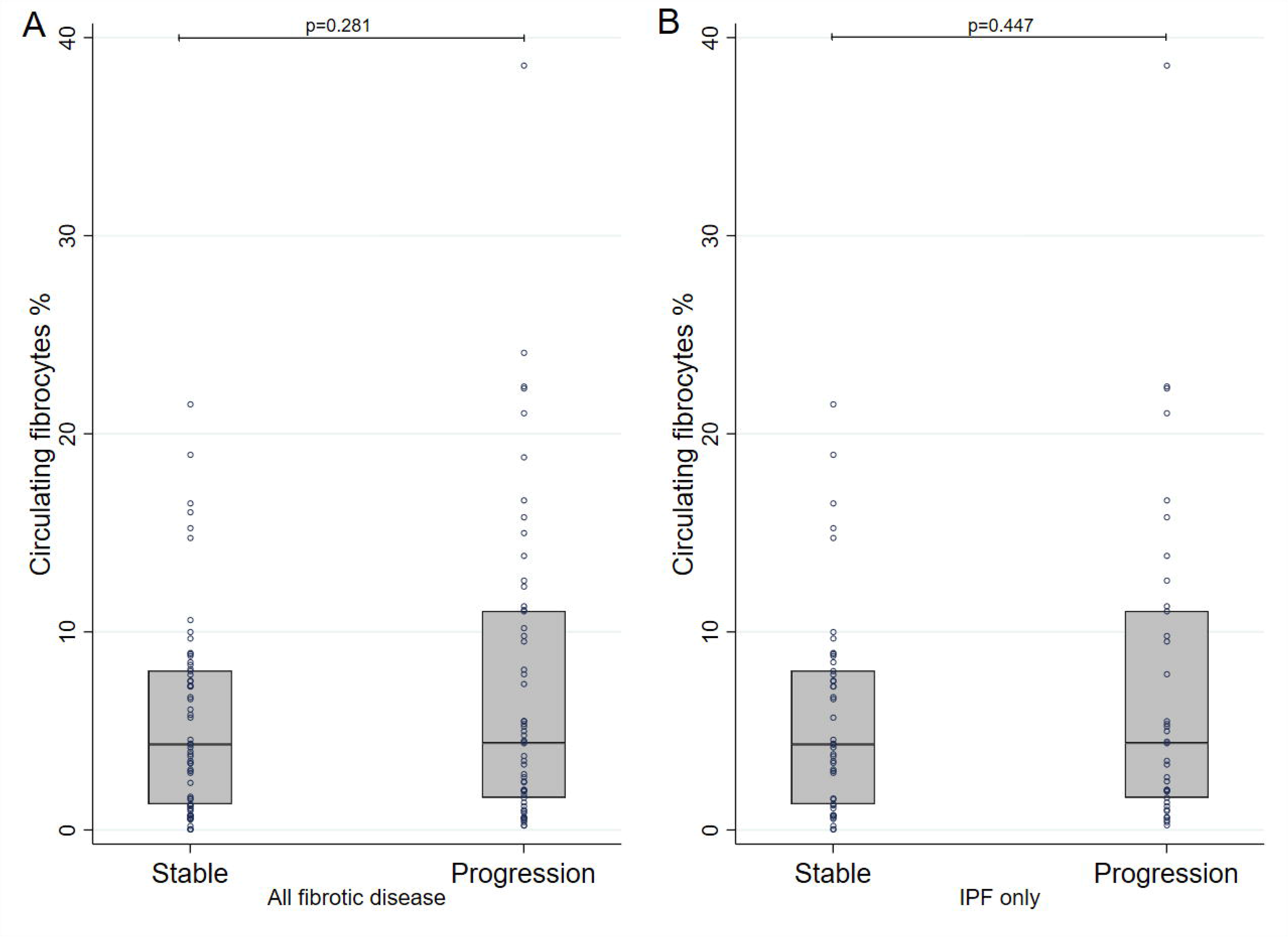
Median circulating fibrocyte level in stable disease and progression. Fibrocyte levels (%) plotted according to disease progression (10% relative decline in FVC or mortality in 12 months), for NSIP and IPF together (A; p=0.281) and IPF alone (B; p=0.447).

No associations between fibrocyte level and lung function metrics were observed in linear models, adjustment for *a priori* confounders of age, gender, smoking and ILD subtype did not alter these findings (Table 4). The effect sizes of fibrocyte level on 12 month delta FVC and delta DLCO values were very small and non-significant (data not shown).

Median fibrocyte levels were not significantly elevated in participants with pulmonary fibrosis whose disease progressed within 12 months (4.9% IQR 2.0%-10.1%), compared with those having stable disease (3.7% IQR 1.1%-8.1% p=0.281, Figure 4). Similar results were observed in IPF diagnosed participants alone (p=0.447).

## Discussion

Using prospectively sampled participants recruited into a well-characterised, multicentre, clinical observational study, we provide data that support the findings of smaller studies but add caution to the clinical utility of circulating fibrocyte levels. In IPF specifically, we demonstrate that a circulating fibrocyte level of 2.22% or over was associated with a 2.5-fold greater risk of mortality. No associations were observed between fibrocyte level and lung function change, or short term disease progression, which suggests circulating fibrocytes offer sensitivity for poor clinical outcomes but poor specificity in distinguishing severity of pulmonary fibrosis.

We demonstrate that variability across participant groups may limit diagnostic potential, and we were unable to reproduce the prognostic value of a 5% threshold previously reported by Moeller *et al*. This discrepancy is likely due to the inclusion of acute exacerbations of IPF in the historic cohort, whilst PROFILE participants were sampled during stability. The PROFILE cohort was used to derive a empirical threshold of 2.22%, which was associated with mortality within the cohort and overall. Steroid therapy for immunosuppression is considered to inhibit differentiation of fibroblasts;^10^ adjustment for a priori confounders in Moeller *et al*. data, particularly steroid immunosuppression, resulted in a wide confidence interval and a non-significant association. Pooling IPF cohorts in a multilevel model provides the largest evaluated sample of fibrocyte data in IPF, and demonstrates a significant association with mortality in adjusted analysis. Overall, these findings support the hypothesis that fibrocytes are associated with a poor prognosis and may play a pathogenic role in progressive pulmonary fibrosis. Further study is required to evaluate the association with prognosis in well-powered cohorts of non-IPF interstitial lung disease.

Elevated levels of circulating fibrocytes have been observed in IPF compared with healthy controls.^6 11-13^ We observe variable levels in all groups including those considered healthy, which is limited by sample size. No difference in fibrocyte levels were observed according to 12month disease progression, and Kaplan-Meier curves demonstrated similar survival estimates within the first year regardless of whether fibrocyte level was greater than the optimal threshold. We find no evidence of circulating fibrocyte dynamics over six months, or a relationship to lung function at baseline or longitudinally. The lack of an observable relationship with lung function or disease activity directly contrasts with published findings from a small sample of IPF patients.^12^ Correlations previously observed in RA-ILD are similarly limited by small numbers of samples.^14^ The present findings suggest that circulating fibrocytes may not provide a specific measure of pulmonary fibrosis disease activity, and may be elevated in underlying comorbidities not captured within the dataset, such as hypertension.^15^

We followed established methods to process and measure fibrocytes, however gating strategies based on CD45/collagen-I have been found to include some contamination of granulocytes,^11^ and the differentiation stage of circulating fibrocytes from precursor monocytes may reflect discrete functional subsets.^16–18^ Ongoing efforts to characterise lung-specific fibrocytes may overcome limitations of contamination and offer further insight into the prognostic potential of specific subtypes of fibroblast precursor cells.^11–19^ Periostin is involved in differentiation of lung specific fibroblasts and associations with disease progression have been observed in lung fibrosis.^20–22^ Further studies to distinguish the precise cellular lineages of circulating fibrocytes are required.

Several limitations were present in these data. A total of 51 individuals had complete fibrocyte data at both baseline and six months, and limited molecular profiling assays for healthy participants prevented further distinction based on functional subgroups. We tested longitudinal differences in fibrocytes where complete, finding minimal change and subsequent rationale for analysis of first chronological instance of fibrocyte level. Minimal change over time in disease groups adds further support that circulating fibrocytes may not be a useful metric of short-term fibrotic disease severity, whilst missingness reflects logistical difficulties in processing and measuring fibrocytes from clinical samples. We tested associatons independently and in pooled analyses with historic data, measured using a consistent strategy.^6^ Whilst we account for steroid immunosuppression therapy across both cohorts, PROFILE participants were naïve to anti-fibrotics during the study and comparable with the historic cohort in this context of disease management. Further study is required to evaluate circulating fibrocytes as a biomarker for clinical response to anti-fibrotic therapy. Recently nintedanib, a licensed treatment for IPF, has been demonstrated to attenuate fibrocyte migration and fibroblast differentiation in vitro and in preclinical animal models.^4 21 23^ Clinical studies determining whether nintedanib affects circulating fibrocytes levels would address such a possibility.

We performed analysis in a prospective observational sample, naïve to anti-fibrotic therapy and largely representative of the wider PROFILE cohort, providing robust estimates for supporting prognosis in IPF. We refine findings from previous study to support prognosis in stable IPF and pool data to provide greater power for evidence of an association with overall mortality in adjusted analysis. Further, we provide robust evidence that contrasts the correlations with lung function decline observed in smaller studies of interstitial lung disease ^12 14^. To our knowledge, these results represent the largest prospective IPF sample of circulating fibrocytes studied and are applicable to real-world incidence of individuals with pulmonary fibrosis.

The value of circulating fibrocytes in prognosticating IPF is limited due to the technical challenges measuring fibrocytes and lack of dynamic change or association with short term disease behaviour. However further characterisation of tissue-specific fibrocytes within prospective interventional clinical trials may offer more accurate assessment of their potential as biomarkers of disease activity.

## Data Availability

Data is available from the Study Steering Committee at reasonable request

